# Associations of arterial thickness, stiffness and blood pressure with brain morphology in early adolescence: A prospective population-based study

**DOI:** 10.1101/2023.10.24.23297502

**Authors:** Serena Defina, Carolina C.V. Silva, Charlotte A.M. Cecil, Henning Tiemeier, Janine F. Felix, Ryan L. Mutzel, Vincent W.V. Jaddoe

## Abstract

**Background:** Arterial wall thickness and stiffness and high blood pressure have been repeatedly associated with poorer brain health. However, previous studies largely focused on mid- or late-life stages. It is unknown whether any arterial health-related brain changes may be observable already in adolescence.

**Methods:** We examined whether *(i)* carotid intima-media thickness, *(ii)* carotid distensibility and *(iii)* systolic and diastolic blood pressure, measured at age 10, were associated with brain volumes and/or white matter microstructure (i.e., fractional anisotropy and mean diffusivity) at age 14. In addition to cross-sectional analyses, we explored associations with longitudinal change in each brain outcome from 10 to 14 years. Analyses were based on 5341 children from the Generation R Study.

**Results:** Higher diastolic blood pressure was associated with lower total brain (*β* [95%CI] = -0.04 [- 0.07;-0.01]) and grey matter volumes (*β* [95%CI] = -0.04 [-0.07;-0.01]) at age 14 years, with stronger associations in higher diastolic blood pressure ranges. Similar associations emerged between systolic blood pressure and brain volumes but these were no longer significant after adjusting for birthweight. No associations were observed between blood pressure and white matter microstructure, nor between carotid intima-media thickness or distensibility and brain morphology.

**Conclusions:** Arterial blood pressure, but not intima-media thickness and distensibility, is associated with structural neuroimaging markers in early adolescence. Volumetric measures may be more sensitive to these early arterial health differences compared to microstructural properties of the white matter, but further studies are needed to confirm these results and assess potential causal mechanisms.

Graphic Abstract

## 1 Introduction

While cardiovascular disease remains the leading cause of death worldwide, the global burden of brain disease is rapidly increasing and is often associated with the same risk factors, as highlighted by the latest statistical update from the American Heart Association^1^. Indeed, known cardiovascular risk factors such as hypertension^2-4^, arterial stiffness^5-7^ and atherosclerosis^8-10^, have been consistently associated with the pathogenesis of cerebrovascular disease (e.g., stroke and white matter lesions) and cognitive decline in the elderly.

Accumulating evidence further suggests that more subtle vascular health–related brain changes may occur in younger adults, far before evident injury events, and even when the overall vascular burden is low. For example, the Framingham Third-Generation Cohort Study (FHS-G3) investigators reported cross-sectional associations of arterial stiffness with lower grey-matter density and white-matter microstructural integrity, assessed with diffusion tensor imaging (DTI), in the fifth decade of life^11, 12^. Higher arterial stiffness was also associated with poorer processing speed and executive function, and with larger lateral ventricular volumes in younger adults (30–45 years) from the FHS-G3^13^. Moreover, in a group of 45-year-old adults from the Coronary Artery Risk Development in Young Adults (CARDIA) Study, higher carotid intima–media thickness (cIMT), a subclinical marker of atherosclerosis, was associated with reduced cerebral blood flow and with poorer cognitive function five years later^14, 15^. In the same cohort, midlife hypertension^16^ and increasing blood pressure trajectories from young adulthood to middle age^17^ were associated with lower cerebral perfusion and volumetric markers of poor white- and grey-matter health at age 50. Blood pressure was also linearly associated with reduced grey-matter volumes and white matter microstructural integrity at age 40 in the FHS-G3 sample^18^ and with grey matter and white matter lesion volume before the age of 45 in the UK Biobank^19^.

Although analogous neuroimaging studies in children are lacking, there is evidence linking elevated blood pressure with cognitive performance already in childhood^20, 21^. Further, related cardiovascular risk factors, such as adiposity^22, 23^ and diabetes^24^ have been shown to associate with brain structural changes in children and adolescents. However, thus far very little attention has been paid to the potential impact of vascular health on the brain during development, despite evidence that arterial stiffening and thickening, and blood pressure dysregulation begin very early in life^25-27^. From a clinical standpoint, characterizing these associations is a fundamental step in the development of early intervention strategies that may prevent abnormal brain development before irreversible structural damage occurs.

Using data from the Generation R Study, a large population-based birth cohort, we investigated how arterial thickness and distensibility, as well as systolic / diastolic blood pressure (SBP / DBP), measured at age 10 associate with brain morphology and microstructure extracted from MRI scans at age 10 and 14 years.

This study was pre-registered (https://osf.io/ryc7e). Briefly, we hypothesized that: 1) higher cIMT, 2) lower distensibility and 3) higher SBP / DBP would associate with *a)* lower total brain volume, *b)* lower grey-matter volume, *c)* lower global fractional anisotropy and/or *d)* higher mean diffusivity.

## 2 Methods

### 2.1 Study population

The study uses data from the Generation R Study, an ongoing population-based prospective cohort study based in the city of Rotterdam, the Netherlands. A detailed cohort description is provided elsewhere^28^. In summary, the cohort included 9749 children born between April 2002 and January 2006. The analytical sample consisted of 5341 singletons who participated in the baseline (i.e., 10-year) visit and had no siblings in the sample (*Figure S1*). Of these children, 2054 had complete structural MRI data and 2308 had complete DTI data. Missing values in all variables of interest were imputed by random-forest multiple imputation.

#### Ethical standards

The study conforms with the World Medical Association Declaration of Helsinki (2013)^29^. Written informed consent was obtained from parents. The medical ethical committee of Erasmus MC, University Medical Center Rotterdam approved the study.

### 2.2 Arterial health

At the age of 10 years, carotid artery ultrasound was performed using the Logiq E9 device (GE Medical Systems, Wauwatosa, Wisconsin) while blood pressure was simultaneously assessed at the right brachial artery. Children were in supine position, with their head tilted slightly away from the transducer. The common carotid artery was identified in a longitudinal plane, ∼10 mm proximal from the carotid bifurcation. Each common carotid artery was measured three times, resulting in six recordings that ideally included multiple heart cycles.

#### Carotid intima–media thickness

For each ultrasound recording, and at all R waves of the simultaneous electrocardiogram, cIMT was computed at the far wall as the average distance between lumen-intima and media-adventitia borders. Average cIMT of all frames of the acquired image sequence was then computed. Analyses were performed offline and semiautomatically, using the application Carotid Studio (Cardiovascular Suite; Quipu srl, Pisa, Italy). Overall mean cIMT (millimeters) was standardized using a z-transformation (i.e., (value – sample mean) / sample SD).

#### Carotid distensibility

The *c* coefficient, i.e., the relative change in lumen area during systole for a given peripheral pressure change was calculated as the difference between the maximal (diastolic) and minimal (systolic) lumen diameter of the carotid artery. Lumen diameter was computed as the average distance between the far and near media-adventitia interfaces for each frame of the acquired image sequence. Per recording, average distension and diameter values were used to compute the average carotid distensibility. Overall mean carotid distensibility (kPa^−1^× 10^−3^) was standardized using a z-transformation.

#### Blood pressure

SBP and DBP were measured at the right brachial artery, four times with 1-min intervals, using the validated automatic sphygmomanometer Datascope Accutorr PlusTM (Paramus, New Jersey, USA). SBP and DBP (mm Hg) were determined by excluding the first measurement and averaging the other measurements, and were standardized using a z-transformation.

### 2.3 Brain imaging

At the ages of 10 and 14 years, participants visited the Generation R research center at Erasmus MC – Sophia Children’s Hospital, where brain Magnetic Resonance Images (MRI) were acquired using a single, dedicated 3-Tesla scanner (General Electric MR750w, Milwaukee, WI, USA) with an eight-channel head coil. To minimize head motion, participants were familiarized with the scanner environment using a mock scanner^30^.

#### Brain volume

High resolution T1-weighted images were obtained with an inversion recovery fast-spoiled gradient recalled sequence (parameters: TR = 8.77 ms, TE = 3.4 ms, TI = 600 ms, flip angle = 10°, FOV = 220×220mm, acquisition matrix = 220×220, slice thickness = 1mm, number of slices = 230, voxel size = 1×1×1mm, ARC Acceleration = 2). Images were processed using FreeSurfer 6.0^31^. The technical details of these procedures are described elsewhere^32^. In brief, this included removal of the non-brain tissue, segmentation of white and grey matter structures, tessellation of the grey-white matter boundary, topology correction and surface deformation to identify the cortical grey-white matter and the grey-cerebrospinal fluid boundary. Reconstructions were visually inspected and those with insufficient quality were further excluded^32^.

Global metrics of volume, i.e., total brain (TBV) and total grey matter volume (GMV), as well as specific subcortical structures’ volumes were extracted. Brain metrics were be standardized using a z-transformation.

#### White matter microstructure

DTI data were obtained using an echo-planar sequence with three b = 0 scans and 35 diffusion-weighted images (b = 1000 s/mm2). The following parameters were used: TR = 12.5 ms, TE = 72.8 ms, FOV = 240×240 mm, acquisition matrix = 120×120, slice thickness = 2 mm, number of slices = 65. Images were pre-processed using <u>FSL 6.0.1</u>. Briefly, non-brain tissue was removed, images were corrected for eddy current-induced distortions and minor head motion using ‘eddy’, and the diffusion gradient table was rotated accordingly. A diffusion tensor was fit at each voxel using a weighted least squares method, and common scalar metrics including global fractional anisotropy (FA) and mean diffusivity (MD) were computed. FA describes the degree to which water diffuses preferentially along one direction (e.g., along a bundle of myelinated axons) and is sensitive to microstructural changes. MD describes the average diffusion in all directions. White matter tracts, were also delineated using fully-automated probabilistic fiber tractography as implemented in FSL AutoPtx^33^. Average FA and MD were calculated for each tract. Global and tract-specific FA and MD values were standardized using a z-transformation. Image quality was assessed by manual and automated inspection^34^.

### 2.4 Covariates

Information on the maternal age and child ethnic background (based on parental country of origin and dichotomized into European vs. non-European) was collected by questionnaire at enrollment. Date of birth and child sex, weight and gestational age were recorded at birth. Both caregivers reported on their highest completed educational level when children were 6 years old, and these reports were combined into a single “parental education” score. Child height (in m) and weight (in kg) were measured during the 10-years visit, and used to compute body mass index (BMI) z-scores.

### 2.5 Statistical analysis

Analyses were conducted using R version 4.2.0^35^. All scripts are <u>publicly available</u>.

#### Imputation

Missing data in all exposures, outcomes and covariates were imputed by random-forest multiple imputation^36^, using 20 imputed datasets, 10 trees and 40 iterations, as implemented by the mice R package^37^. Details of the imputation model and quality are provided in *Supplementary materials* (*Methods S1* and *Table S1*).

#### Main analyses

All models were fit in each imputed dataset, and pooled across imputations using Rubin’s rules^38^. For each exposure of interest (i.e., carotid IMT and distensibility, SBP and DBP), four multiple linear regressions were performed including *a)* TBV, *b)* GMV, *c)* global FA and *d)* global MD, measured at age 14, as dependent variable. We ran a “base model” adjusting for child sex, height, age at MRI assessment and age gap between clinical and MRI assessments, and a “confounder model”, which additionally included child ethnicity, BMI z-score, parental education and maternal age. Covariates were identified based on the graphical criteria for confounding (*Figure S2*).

To minimize false positive findings due to multiple testing (*k*=16), false discovery rate (FDR) correction^39^ was applied to all p-values.

Non-linear terms for each arterial health exposure (i.e., natural splines) were retained in the model when they significantly improved its fit (see *Methods S2*).

#### Exploratory follow-up analyses

We further assessed associations between each exposure and the longitudinal change in each neuroimaging marker from age 10 to 14 years, using linear mixed-effects models with a random intercept per subject (see *Methods S3*). Note that brain outcomes were not standardized for these analyses to prevent incorrect estimation of the correlation structure and likelihood values.

To further characterize the regional specificity of these effects, we assessed associations with: total intracranial, cerebro-spinal fluid, white matter, cortical and subcortical grey matter volumes, as well as subcortical regional volumes (Accumbens, Amygdala, Caudate, Hippocampus, Pallidum, Putamen, Thalamus); white-matter tracts FA and MD (Cingulate gyrus, Cortico-spinal tract, Uncinate fasciculus, Inferior & Superior longitudinal fasciculus, Major & Minor forceps), and vertex-wise cortical thickness (see *Methods S4*) at age 14. These latter analyses were further adjusted for total intracranial volume.

Finally, since sex differences and premature birth have been implicated in the associations of interest^40, 41^, we *a)* investigated effect modification by sex and *b)* additionally adjusted the main models for birthweight and gestational age at birth.

#### Sensitivity analyses

To assess the impact and adequacy of our sample selection and imputation procedure, we ran all analyses in the full cohort (*n*=9749) and in the subsample with complete outcome data (*n*=2054-2308).

## 3 Results

### Participants characteristics

Sample descriptives are displayed in *Table 1* (see also *Tables S1-S2*).

**Table 1.**
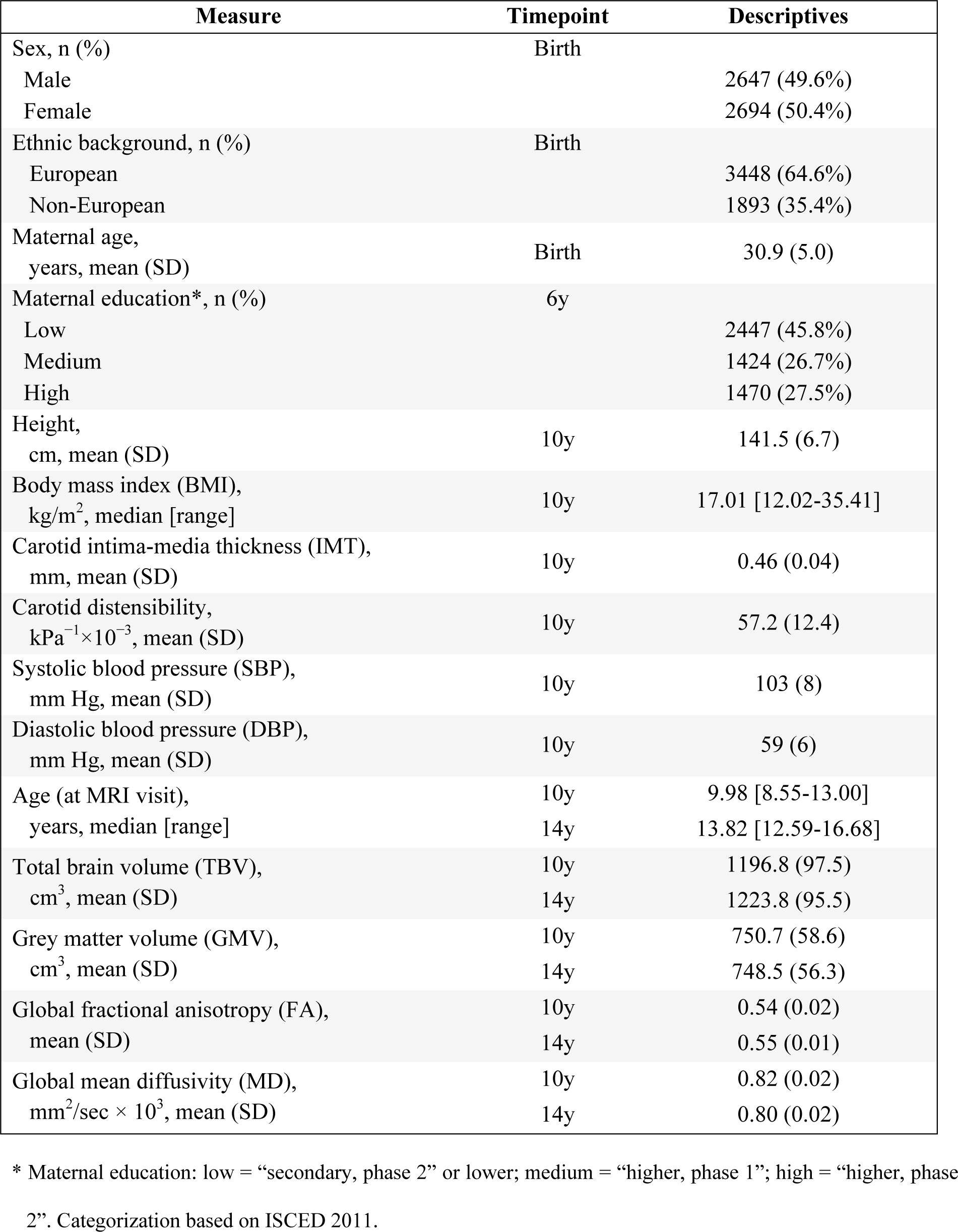
Sample descriptives.

| Measure | Timepoint | Descriptives |
| --- | --- | --- |
| Sex, n (%) | Birth |  |
| Male |  | 2647 (49.6%) |
| Female |  | 2694 (50.4%) |
| Ethnic background, n (%) | Birth |  |
| European |  | 3448 (64.6%) |
| Non-European |  | 1893 (35.4%) |
| Maternal age, years, mean (SD) | Birth | 30.9 (5.0) |
| Maternal education*, n (%) | 6y |  |
| Low |  | 2447 (45.8%) |
| Medium |  | 1424 (26.7%) |
| High |  | 1470 (27.5%) |
| Height, cm, mean (SD) | 10y | 141.5 (6.7) |
| Body mass index (BMI), kg/m<sup>2</sup>, median [range] | 10y | 17.01 [12.02-35.41] |
| Carotid intima-media thickness (IMT), mm, mean (SD) | 10y | 0.46 (0.04) |
| Carotid distensibility, kPa<sup>-1</sup>×10<sup>-3</sup>, mean (SD) | 10y | 57.2 (12.4) |
| Systolic blood pressure (SBP), mm Hg, mean (SD) | 10y | 103 (8) |
| Diastolic blood pressure (DBP), mm Hg, mean (SD) | 10y | 59 (6) |
| Age (at MRI visit), years, median [range] | 10y<br>14y | 9.98 [8.55-13.00]<br>13.82 [12.59-16.68] |
| Total brain volume (TBV), cm<sup>3</sup>, mean (SD) | 10y<br>14y | 1196.8 (97.5)<br>1223.8 (95.5) |
| Grey matter volume (GMV), cm<sup>3</sup>, mean (SD) | 10y<br>14y | 750.7 (58.6)<br>748.5 (56.3) |
| Global fractional anisotropy (FA), mean (SD) | 10y<br>14y | 0.54 (0.02)<br>0.55 (0.01) |
| Global mean diffusivity (MD), mm<sup>2</sup>/sec × 10<sup>3</sup>, mean (SD) | 10y<br>14y | 0.82 (0.02)<br>0.80 (0.02) |

\* Maternal education: low = “secondary, phase 2” or lower; medium = “higher, phase 1”; high = “higher, phase 2”. Categorization based on ISCED 2011.

### Main analyses

An overview of the main results is presented in *Figure 1*. Complete model outputs are reported in supplementary materials (*Table S3*). Neither carotid IMT nor distensibility were significantly associated with any brain outcome (*Figure 1A-B*, *Table S3*). After adjustment for potential confounders, SBP was (cross-sectionally) associated with lower TBV (*β* [95%CI] = -0.04 [- 0.07;-0.01], *P_FDR_* = .030) and GMV (*β* [95%CI] = -0.05 [-0.08;-0.01], *P_FDR_* = .030) but not with either FA or MD (*Figure 1C*, *Table S3*). Analogously, DBP was (cross-sectionally) associated with TBV (*β* [95%CI] = -0.04 [-0.07;-0.01], *P_FDR_* = .022) and GMV (*β* [95%CI] = -0.04 [-0.07;-0.01], *P_FDR_* = .022),but not FA or MD (*Figure 1D*, *Table S3*).

**Figure 1.** Arterial thickness, stiffness and blood pressure and brain morphology at 14 years. For each exposure of interest: **A.** intima-media thickness (IMT); **B.** carotid distensibility; **C.** Systolic blood pressure (SBP); and **D.** Diastolic blood pressure (DBP), the standardized association estimates and their 95% confidence intervals are displayed on the x-axis for each outcome (total brain volume (TBV) in blue; grey matter volume (GMV) in grey; global fractional anisotropy (FA) in red; and mean diffusivity (MD) in orange). The corresponding FDR-corrected P-values are also reported.

We found evidence of a non-linear relationship between SBP and white matter microstructure (*P*=.032 for FA; *P*=.023 for MD) and between DBP and brain volumes (*P*=.012 for TBV; *P*=.006 for GMV). However, these departures from linearity were small, as shown in *Figure 2* and *Figure S3*.

**Figure 2.** Non-linear relationships between blood pressure and brain morphology at 14 years. The linear and non-linear relationship between SBP and **(A)** FA; and **(C)** MD; and between DBP and **(B)** TBV; and **(D)** GMV. Linear associations (black dashed line) were pooled across datasets (and correspond to the estimates presented in Figure 1), while non-linear associations (green continuous lines) were fit in each imputed dataset individually using natural splines with 4 knots. The gray vertical shadows also mark the –2.5 and +2.5 SD cutoffs of each exposure distribution.

### Exploratory follow-up analyses

In our exploratory longitudinal models, we did not find any significant interaction between arterial health markers and age at MRI measurement (*Table S4*; *Figure 3*). However, when interaction terms were excluded from the models, we could further confirm a negative main effect of SBP and DBP on total brain and grey matter volumes measured at 10 and 14 years (*Table S5*). Specifically, while TBV increased with age (*b* [95%CI] = 7.0 [6.4; 7.7] cm^3^ per year) each SD increase in SBP was associated with -3.6 [-6.4; -0.8] cm^3^ (*P_FDR_* =.048) and each SD increase in DBP was associated with -3.4 [-6.1; -0.7] cm^3^ (*P_FDR_* =.040) TBV. Conversely, GMV showed a slight decline over our age range (*b* [95%CI] = -0.6 [-1.0; -0.2] cm^3^ per year) while each SD increase in SBP was associated with -2.4 [-4.1; -0.7] cm^3^ (*P_FDR_* =.042) and each SD increase in DBP was associated with -2.2 [-3.8; -0.6] cm^3^ (*P_FDR_* =.039) GMV.

**Figure 3.** Structural brain changes from 10 to 14 years. The longitudinal change in total brain **(A, B)** and grey matter volume **(C, D)** is represented for children with “high” levels of exposure (i.e., > 1 SD above the mean in systolic or diastolic blood pressure; in red), exposure values in the intermediate rage (i.e., between –1 and 1 SD around the mean; in green) or low levels of the exposure (i.e., < –1 SD below the mean; in blue). The distribution of age at MRI measurement is also depicted in grey on the bottom of each graph.

Associations were largely homogenous between sexes (*Figure S4*, *Table S6*) and did not seem to be explained by differences in cranium size, nor by a significant increase in cerebro-spinal fluid volumes (*Figure S5, Table S7*). There were no significant associations between either exposure and total or region-specific subcortical volumes (*Figure S5, Table S8*) nor with white matter tracts FA and MD (*Figures S6-S7*, *Tables S9-S10*). SBP was significantly associated with total cortical volume (*β* [95%CI] = -0.05 [-0.08;-0.01], *P_FDR_* = .036; see *Figure S5* and *Table S7*), but no local associations with cortical thickness emerged from the vertex-wise analyses (*Table S11*).

Notably, the associations between SBP and total/grey matter volumes were significantly attenuated after adjusting for birth-related covariates (TBV: *β* [95%CI] = -0.03 [-0.06; 0.01], *P_FDR_* = .230; GMV: *β* [95%CI] = -0.03 [-0.06; 0.00], *P_FDR_*= .229; see *Table S12*).

### Sensitivity analyses

Restricting the analyses to participants with complete outcome data (N = 2054 for structural MRI and 2308 for DTI) did not substantively change the reported findings, besides a slight increase in effect sizes for both SBP (TBV: *β* [95%CI] = -0.05 [-0.09;-0.01], *P_FDR_* = .016; GMV: *β* [95%CI] = -0.06 [-0.09;-0.02], *P_FDR_*= .013) and DBP (TBV and GMV: *β* [95%CI] = -0.05 [-0.09;-0.02], *P_FDR_* = .012; see *Tables S13-S14*).

## 4 Discussion

In this population-based prospective cohort study, we observed that systolic and diastolic blood pressure at age 10 years were associated with lower total and grey matter volumes at age 10 and 14 years. The associations between systolic blood pressure and brain volumes were partially explained by birth-related confounders (particularly birthweight). No significant associations were observed for carotid intima-media thickness or distensibility, nor for white matter microstructural markers.

### Interpretation of main findings

To the best of our knowledge, this is the first study to establish a link between high blood pressure and brain volume reduction in the general pediatric population. Together with previous pediatric neuroimaging studies focusing mainly on adiposity^22, 23, 42^, our findings confirm the idea that, already at school age and within subclinical ranges, an adverse cardiovascular profile may negatively impact brain development.

We could not find evidence for a significant role of arterial wall thickness or stiffness on the adolescent brain, as we had expected based on adult reports^11, 14^. This might have been due to insufficient sensitivity of ultrasound measures and/or lack of variability in these markers in healthy pediatric population.

In line with our findings, several previous studies reported a negative association between (diastolic) blood pressure and cognitive function in children and adolescents^20, 21, 43, 44^. The biological substrate underlying these associations, however, is less studied. Interestingly, a few transcranial Doppler ultrasound studies demonstrated blunted cerebrovascular reactivity in hypertensive children^20, 45^, however, these were conducted in small samples and suffer from wide inter-observer measurement variability. We corroborated and extended these findings using more stable structural MRI markers, and found that morphological brain outcomes (volumetric measures) may be already sensitive to early increase in arterial blood pressure. This was not true for microstructural properties of the white matter, which was unexpected, as adult studies typically point to impaired white matter integrity as an early marker of neurovascular pathology^18^. However, it is important to note that both macroscopic and microstructural changes reported in older populations may reflect different processes (e.g., cellular atrophy and white matter lesions) compared to earlier developmental windows, where the same neuroimaging markers may underly, for instance, grey and white matter maturation. Indeed, while grey matter maturation tends to peak already in early adolescence, white matter growth typically doesn’t peak until mid-adulthood, which could partly explain the observed pattern of results^46^.

### Biological mechanisms

Several potential mechanisms could explain the reported associations. For example, it was been proposed that high blood pressure could cause damage to the neurovasculature and result in reduced cerebral blood perfusion, leading to suboptimal oxygen and nutritional supplies, and potentially altering ongoing neurodevelopmental processes^44^. Additionally, alterations in immune and hypothalamic-pituitary-adrenal axis functioning resulting from chronically high blood pressure could trigger neuroinflammation, further impairing brain development^47^.

### Clinical relevance

Over the past decades, the prevalence of high blood pressure among children and adolescents has increased dramatically, in concert with the global epidemic of obesity^48^. Elevated blood pressure in childhood is known to track into adulthood and increase the risk of cardiovascular disease^49^. In our 10-year-old sample, each 6 mmHg increase in diastolic blood pressure (i.e., 1 SD) was followed by a 3.6 cm^3^ reduction in total brain volume, and a 2.3 cm^3^ reduction in grey matter volume four years later. In our longitudinal follow-up analysis, we could further show how this corresponded to about one third of the estimated yearly change in TBV. These results are important from a developmental perspective, since they suggest that early detection and prevention of elevated blood pressure may be relevant not only for long-term cardiovascular health but also for structural brain development. However, whether these associations are clinically relevant (e.g., in terms of cognitive performance or mental health symptomatology) or whether they will evolve into long-term alteration of brain health, needs to be further studied. Nevertheless, pediatric screening for high blood pressure may be a relatively easy measure to implement to monitor and potentially decrease the risk of future brain disease and cognitive impairment.

### Strengths and limitations

This study used prospectively measured data from a large population-based sample. Detailed and objective measures of both arterial and brain health were available and selection bias due to nonresponse was addressed thoroughly using multiple imputation and sensitivity analyses. However, it is important to note that, although we adjusted the analyses for several sociodemographic and lifestyle factors known to influence the associations, residual confounding, for example by genetic predisposition, exposure to stress, nutritional intake or physical activity, may still be present. Additionally, note that, for practical reasons, blood pressure was measured in supine position in this study, as opposed to more conventional sitting measures. Further studies are underlying mechanisms and/or modifiable factors that could explain or attenuate these associations.

### Perspectives

High blood pressure in childhood was associated with suboptimal brain development already in early adolescence, particularly with respect to reduced total brain and grey matter volumes. Pediatric screening for high blood pressure may thus be relevant not only for the prevention of long-term cardiovascular health problems, but also for structural brain development. Identifying such early biomarkers of neurovascular health entails important implications for future clinical and public heath priorities, especially considering the established relationship between these markers and later prognosis of cerebrovascular disease and dementia.

### Novelty and Relevance

- <u>What Is New</u>? Higher blood pressure is related to smaller brain volumes already during early adolescence.
- <u>What Is Relevant</u>? This is the first study to show that higher blood pressure, even within subclinical ranges, may have negative consequences on adolescent brain development.
- <u>Clinical/Pathophysiological Implications</u>? Among the many factors that can influence a healthy cognitive and psychological development, blood pressure may be an underappreciated one.

## Data Availability

SD had full access to all the data in the study and takes responsibility for its integrity and the data analysis. Data from the Generation R Study are not publicly available as the consent given by the participants does not allow for storage of individual-level data in repositories or journals. Researchers who want to access the dataset for replication should submit a request to the director of the Generation R Study. Data access is subject to local, national and European rules and regulations. All scripts employed in data preparation and analysis are publicly available.

## Non-standard Abbreviations and Acronyms

BMI: Body mass index
cIMT: Carotid intima–media thickness
CARDIA: Coronary Artery Risk Development in Young Adults
DTI: Diffusion tensor imaging
FDR: False discovery rate
FA: Fractional anisotropy
FHS-G3: Framingham Third-Generation Cohort Study
GMV: Grey matter volume
MRI: Magnetic Resonance Images
MD: Mean diffusivity
SBP / DBP: Systolic / diastolic blood pressure
TBV: Total brain volume

## Acknowledgments

The Generation R Study is conducted by Erasmus MC, University Medical Center Rotterdam in close collaboration with the School of Law and Faculty of Social Sciences of the Erasmus University Rotterdam, the Municipal Health Service Rotterdam area, Rotterdam, the Rotterdam Homecare Foundation, Rotterdam and the Stichting Trombosedienst & Artsenlaboratorium Rijnmond (STAR-MDC), Rotterdam. We gratefully acknowledge the contribution of children and parents, general practitioners, hospitals, midwives and pharmacies in Rotterdam.

Author contributions are presented according to the CRediT (Contributor Roles Taxonomy). S.D.: Conceptualisation, methodology, formal analysis, software, data curation, visualization, writing - original draft, writing -review & editing, project administration. C.S.: Conceptualisation, writing - review & editing. C.C.: Writing -review & editing, supervision, funding acquisition. H.T.: Writing - review & editing, supervision. J.F.: Writing -review & editing, supervision, funding acquisition. R.M.: Conceptualisation, methodology, writing -review & editing. V.J.: Conceptualisation, methodology, writing -review & editing, supervision. All authors read and approved the final manuscript.

## 8 References

1. Tsao CW, Aday AW, Almarzooq ZI, Alonso A, Beaton AZ, Bittencourt MS, et al. Heart disease and stroke statistics—2022 update: A report from the american heart association. Circulation. 2022;145:e153–e639

2. Kelly DM, Rothwell PM. Blood pressure and the brain: The neurology of hypertension. Practical Neurology. 2020;20:100

3. Beauchet O, Celle S, Roche F, Bartha R, Montero-Odasso M, Allali G, et al. Blood pressure levels and brain volume reduction: A systematic review and meta-analysis. J Hypertens. 2013;31:1502–1516

4. Alateeq K, Walsh EI, Cherbuin N. Higher blood pressure is associated with greater white matter lesions and brain atrophy: A systematic review with meta-analysis. Journal of Clinical Medicine. 2021;10:637

5. Iulita MF, Noriega de la Colina A, Girouard H. Arterial stiffness, cognitive impairment and dementia: Confounding factor or real risk? Journal of Neurochemistry. 2018;144:527–548

6. Baradaran H, Gupta A. Carotid artery stiffness: Imaging techniques and impact on cerebrovascular disease. Frontiers in Cardiovascular Medicine. 2022;9

7. Badji A, Sabra D, Bherer L, Cohen-Adad J, Girouard H, Gauthier CJ. Arterial stiffness and brain integrity: A review of mri findings. Ageing Research Reviews. 2019;53:100907

8. Wang W, Norby FL, Alonso A, Gottesman RF, Jack CR, Jr., Meyer ML, et al. Association of carotid intima-media thickness with brain mri markers in the atherosclerosis risk in communities neurocognitive study (aric-ncs). Journal of Stroke and Cerebrovascular Diseases. 2022;31

9. Álvarez-Bueno C, Cavero-Redondo I, Bruno RM, Saz-Lara A, Sequí-Dominguez I, Notario-Pacheco B, et al. Intima media thickness and cognitive function among adults: Metanalysis of observational and longitudinal studies. Journal of the American Heart Association. 2022;11:e021760

10. Baradaran H, Gupta A. Brain imaging biomarkers of carotid artery disease. Ann Transl Med. 2020;8:1277–1277

11. Maillard P, Mitchell GF, Himali JJ, Beiser A, Tsao CW, Pase MP, et al. Effects of arterial stiffness on brain integrity in young adults from the framingham heart study. Stroke. 2016;47:1030–1036

12. Maillard P, Mitchell GF, Himali JJ, Beiser A, Fletcher E, Tsao CW, et al. Aortic stiffness, increased white matter free water, and altered microstructural integrity: A continuum of injury. Stroke. 2017;48:1567–1573

13. Pase MP, Himali JJ, Mitchell GF, Beiser A, Maillard P, Tsao C, et al. Association of aortic stiffness with cognition and brain aging in young and middle-aged adults. Hypertension. 2016;67:513–519

14. Cermakova P, Ding J, Meirelles O, Reis J, Religa D, Schreiner PJ, et al. Carotid intima-media thickness and markers of brain health in a biracial middle-aged cohort: Cardia brain mri sub-study. J Gerontol A Biol Sci Med Sci. 2020;75:380–386

15. Al Hazzouri AZ, Vittinghoff E, Sidney S, Reis JP, Jacobs Jr DR, Yaffe K. Intima-media thickness and cognitive function in stroke-free middle-aged adults: Findings from the coronary artery risk development in young adults study. Stroke. 2015;46:2190–2196

16. Launer LJ, Lewis CE, Schreiner PJ, Sidney S, Battapady H, Jacobs DR, et al. Vascular factors and multiple measures of early brain health: Cardia brain mri study. PLoS One. 2015;10:e0122138

17. Hu Y-H, Halstead MR, Bryan RN, Schreiner PJ, Jacobs DR, Jr, Sidney S, et al. Association of early adulthood 25-year blood pressure trajectories with cerebral lesions and brain structure in midlife. JAMA Network Open. 2022;5:e221175–e221175

18. Maillard P, Seshadri S, Beiser A, Himali JJ, Au R, Fletcher E, et al. Effects of systolic blood pressure on white-matter integrity in young adults in the framingham heart study: A cross-sectional study. The Lancet Neurology. 2012;11:1039–1047

19. Alateeq K, Walsh EI, Abhayaratna WP, Cherbuin N. Effects of higher normal blood pressure on brain are detectable before middle-age and differ by sex. Journal of Clinical Medicine. 2022;11:3127

20. Lande MB, Kupferman JC. Blood pressure and cognitive function in children and adolescents. Hypertension. 2019;73:532–540

21. Lamballais S, Sajjad A, Leening MJG, Gaillard R, Franco OH, Mattace-Raso FUS, et al. Association of blood pressure and arterial stiffness with cognition in 2 population-based child and adult cohorts. Journal of the American Heart Association. 2018;7:e009847

22. Silva CCV, Jaddoe VWV, Muetzel RL, Santos S, El Marroun H. Body fat, cardiovascular risk factors and brain structure in school-age children. International Journal of Obesity. 2021;45:2425–2431

23. Steegers C, Blok E, Lamballais S, Jaddoe V, Bernardoni F, Vernooij M, et al. The association between body mass index and brain morphology in children: A population-based study. Brain Struct Funct. 2021;226:787–800

24. Redel JM, DiFrancesco M, Lee GR, Ziv A, Dolan LM, Brady CC, et al. Cerebral blood flow is lower in youth with type 2 diabetes compared to obese controls: A pilot study. Pediatric Diabetes. 2022;23:291–300

25. Song P, Fang Z, Wang H, Cai Y, Rahimi K, Zhu Y, et al. Global and regional prevalence, burden, and risk factors for carotid atherosclerosis: A systematic review, meta-analysis, and modelling study. The Lancet Global Health. 2020;8:e721–e729

26. Rosner B, Cook NR, Daniels S, Falkner B. Childhood blood pressure trends and risk factors for high blood pressure: The nhanes experience 1988–2008. Hypertension. 2013;62:247–254

27. Kruger R, Gafane-Matemane LF, Kagura J. Racial differences of early vascular aging in children and adolescents. Pediatric Nephrology. 2021;36:1087–1108

28. Kooijman MN, Kruithof CJ, van Duijn CM, Duijts L, Franco OH, van Ijzendoorn MH, et al. The generation r study: Design and cohort update 2017. Eur J Epidemiol. 2016;31:1243–1264

29. World medical association declaration of helsinki: Ethical principles for medical research involving human subjects. Jama. 2013;310:2191-2194

30. White T, Muetzel RL, El Marroun H, Blanken LME, Jansen P, Bolhuis K, et al. Paediatric population neuroimaging and the generation r study: The second wave. Eur J Epidemiol. 2018;33:99–125

31. Fischl B. Freesurfer. Neuroimage. 2012;62:774–781

32. Muetzel RL, Mulder RH, Lamballais S, Cortes Hidalgo AP, Jansen P, Güroğlu B, et al. Frequent bullying involvement and brain morphology in children. Frontiers in Psychiatry. 2019;10

33. De Groot M, Vernooij MW, Klein S, Ikram MA, Vos FM, Smith SM, et al. Improving alignment in tract-based spatial statistics: Evaluation and optimization of image registration. Neuroimage. 2013;76:400–411

34. Muetzel RL, Blanken LME, van der Ende J, El Marroun H, Shaw P, Sudre G, et al. Tracking brain development and dimensional psychiatric symptoms in children: A longitudinal population-based neuroimaging study. Am J Psychiatry. 2018;175:54–62

35. R Core Team. R: A language and environment for statistical computing. 2021

36. Shah AD, Bartlett JW, Carpenter J, Nicholas O, Hemingway H. Comparison of random forest and parametric imputation models for imputing missing data using mice: A caliber study. Am J Epidemiol. 2014;179:764–774

37. Buuren S, Groothuis-Oudshoorn C. Mice: Multivariate imputation by chained equations in r. Journal of Statistical Software. 2011;45

38. Rubin DB. Multiple imputation for survey nonresponse. 1987

39. Benjamini Y, Yekutieli D. The control of the false discovery rate in multiple testing under dependency. Annals of statistics. 2001:1165–1188

40. Pasha EP, Birdsill AC, Oleson S, Tanaka H, Haley AP. Associations of carotid arterial compliance and white matter diffusion metrics during midlife: Modulation by sex. Neurobiology of Aging. 2018;66:59–67

41. Gluckman PD, Hanson MA, Cooper C, Thornburg KL. Effect of in utero and early-life conditions on adult health and disease. New England Journal of Medicine. 2008;359:61–73

42. Group BDC. Total and regional brain volumes in a population-based normative sample from 4 to 18 years: The nih mri study of normal brain development. Cerebral Cortex. 2012;22:1–12

43. Dawson AE, Kallash M, Spencer JD, Wilson CS. The pressure’s on: Understanding neurocognitive and psychological associations with pediatric hypertension to inform comprehensive care. Pediatric Nephrology. 2021;36:3869–3883

44. Lucas I, Puteikis K, Sinha MD, Litwin M, Merkevicius K, Azukaitis K, et al. Knowledge gaps and future directions in cognitive functions in children and adolescents with primary arterial hypertension: A systematic review. Front Cardiovasc Med. 2022;9:973793

45. Wong LJ, Kupferman JC, Prohovnik I, Kirkham FJ, Goodman S, Paterno K, et al. Hypertension impairs vascular reactivity in the pediatric brain. Stroke. 2011;42:1834–1838

46. Groeschel S, Vollmer B, King MD, Connelly A. Developmental changes in cerebral grey and white matter volume from infancy to adulthood. International Journal of Developmental Neuroscience. 2010;28:481–489

47. Perrotta M, Lembo G, Carnevale D. The interactions of the immune system and the brain in hypertension. Current Hypertension Reports. 2018;20:7

48. Riley M, Hernandez AK, Kuznia AL. High blood pressure in children and adolescents. Am Fam Physician. 2018;98:486–494

49. Yang L, Magnussen CG, Yang L, Bovet P, Xi B. Elevated blood pressure in childhood or adolescence and cardiovascular outcomes in adulthood. Hypertension. 2020;75:948–955

